# Structural Variants in Linkage Disequilibrium with GWAS-Significant SNPs

**DOI:** 10.1101/2022.12.14.22283482

**Authors:** Hao Liang, Joni Sedillo, Steven J. Schrodi, Akihiro Ikeda

## Abstract

**Summary:** With the recent expansion of structural variant identification in the human genome, understanding the role of these impactful variants in disease architecture is critically important. Currently, a large proportion of genome-wide-significant GWAS SNPs are functionally unresolved, raising the possibility that some of these SNPs are associated with disease through linkage disequilibrium with causal structural variants. Hence, understanding the linkage disequilibrium between newly discovered structural variants and statistically significant SNPs may provide a resource for further investigation into disease-associated regions in the genome. Here we present a resource cataloging structural variant-significant SNP pairs in high linkage disequilibrium.

**Availability:** All data files including those detailing SV and GWAS SNP associations and results of GWAS-SNP-SV pairs are available at https://github.com/hliang-SchrodiLab/SV_SNPs.

---

Large genomic imbalances can significantly disrupt important functional elements in the human genome including chromatin structure, noncoding RNAs, protein-coding sequence, and gene regulation.^1-3^ These changes can substantially alter phenotypes and potentially drive a wide-array of disease and disease-related traits. These changes can therefore drive substantial phenotypic effects, including those effects that impact the risk of disease. Indeed, structural variants (SVs) have been associated with several clinical traits including schizophrenia^4^, cardiometabolic physiology^5^, amyotrophic lateral sclerosis^6^, low density lipoprotein levels^7^, and neurodevelopmental disorders^8^. However, short-read sequencing technology has limited ability to detect SVs.^9^ To address this limitation, a study of 32 human genomes was recently conducted by the Human Genome Structural Variation Consortium (HGSVC) using a combination of long-read PacBio whole genome sequencing and Strand-seq technologies, identifying 107,590 SVs across the genome.^10^ The authors noted that 68% of these SVs were not discovered by short-read sequencing. In our study, we sought to determine which single nucleotide polymorphisms (SNPs), exceeding genome-wide significance levels in genome-wide association studies (GWAS) were in high linkage disequilibrium with newly identified SVs. By doing so, these results generate specific hypotheses concerning SVs as causal variants which could drive correlated SNPs to exhibit strong disease association. Hence, this work can serve as a resource to investigate disease-association at SVs not interrogated in GWAS which may be driving disease signals.

To construct the database of GWAS-significant SNPs (p<5E-08) in high linkage disequilibrium with these new SVs, SNPs were obtained from the GWAS catalog^11^ noting the position (assembly GRch38/hg38) and alleles. SV location and alleles were downloaded from the HGSVC data portal.^12^ To reduce the computational effort, a restriction was placed on the physical distance between GWAS SNPs and SVs prior to calculating linkage disequilibrium. This distance was set to 100kbp flanking the endpoints of the SV. Using the unphased data, linkage disequilibrium was then calculated on the HGSVC samples using the approach described below. GWAS SNP-SV pairs that exceeded a squared correlation coefficient of 0.80 were included in the database.

Perl (v5.32.1) code was used to preprocess original files, including file format conversion, GWAS association extraction and sample counting. Code written in R (v4.1.3) was used to calculate linkage disequilibrium and p-values. Bedtools (v2.30.0) was also used to extract SNPs located within the regions of SVs and their upstream/downstream 100kbp flanking regions.

To calculate the pairwise linkage disequilibrium between each SV and nearby GWAS-significant SNPs, an estimator of the squared correlation coefficient was used on unphased data. The SVs and SNPs studied are biallelic. Denote the pair of alleles segregating at an SV as *A*_1_ and *A*_2_. Similarly denote the pair of alleles segregating at a SNP as *B*_1_ and *B*_2_. Further denote the number of the nine double genotypes in a sample of individuals as:

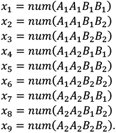

Let 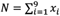.

Setting the numerical values for each individual carrying a specific genotype as

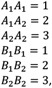

we then define the genotypic squared correlation coefficient (Pearson’s correlation coefficient squared) as

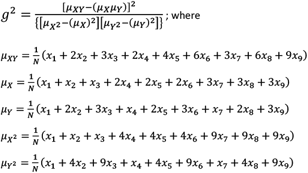

Notably, the value of *g*^2^ is equivalent to the standard metric *r*^2^ under Hardy-Weinberg equilibrium.

Our analysis produced 16,238 GWAS-SNP-SV pairs in high linkage disequilibrium (g^2^ > 0.80) across 2,355 traits from the GWAS catalog within the physical distance window. These SNP-SV pairs were composed of a total of 4,677 unique SVs and 7,831 unique GWAS-significant SNPs. The distribution of numbers of high linkage disequilibrium SNP-SV pairs for each chromosome is shown in **Supplemental Table 1**. To exemplify the utility of this resource, we show the findings for nine GWAS SNPs within the ±100kbp flanking region of SV10995 (insertion/deletion of 384 nucleotides) in the *PLEKHA1/ARMS2/HTRA1* region on chromosome 10q26 (**Figure 1**). Six of these SNPs, rs11200638, rs3793917, rs3750846, rs3750847, rs3750848, and rs10490924 were all previously found to be highly associated with age-related macular degeneration (AMD).^13-15^ Four of these six SNPs were found to be in perfect linkage disequilibrium with SV10995 within the HGSVC sample set and rs11200638 and rs3793917 exhibited a *g*^2^ = 0.94. Outside of AMD, rs61871747 was suggestively associated with cognitive function (p=9.13E-07), rs36212732 was found to significantly correlate with refractive error, and rs61871744 was significantly associated with cataract. SV10995 resides immediately (11bp (gene body) / 431bp(cds)) downstream of *ARMS2*. It is possible that SV10995 affects the expression of ARMS2, which may, in turn, modify the risk for AMD. Interestingly, a different SV^16^ was found to both be associated with AMD and have substantial effects on *ARMS2* mRNA stability.^17^

**Figure 1.**
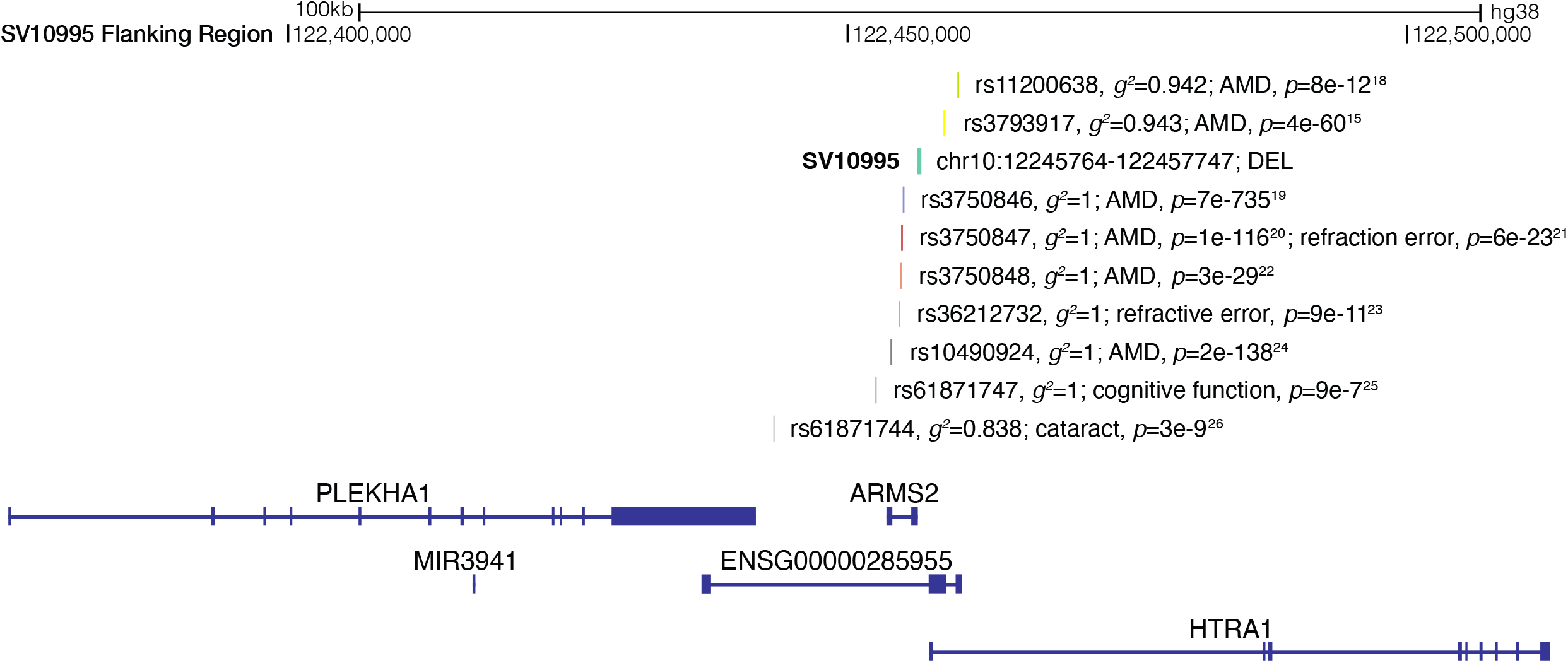
*Example: SV10995* and Significant GWAS SNPs in High Linkage Disequilibrium. Figure 1 shows the region from chr10:122357364-122557747 (hg38 assembly) showing SV10995 (insertion/deletion structural variant), the ten SNPs associated with disease traits from the GWAS Catalog, the squared correlation (*g*^2^) between each SNP and SV10995, and the p-value reported in the respective publication for each SNP. This information is displayed positionally in the context of *PLEKHA1*, the microRNA *MIR3941*, ENSG00000285955, *ARMS2*, and *HTRA1*.

Through the creation of this repository of disease-associated SNPs that are highly correlated with newly discovered SVs, this archive can serve as an important resource for fine mapping causal variants in medically important traits.

## Data Availability

All data files including those detailing SV and GWAS SNP associations and results of GWAS SNP-SV pairs are available at https://github.com/hliang-SchrodiLab/SV_SNPs

---

**Supplemental Table 1.** The number of genome-wide significant SNPs in high linkage disequilibrium (*g*^2^ > 0.80) with a structural variant listed by chromosome.

| Chromosome | SNP-SV Pairs |
| --- | --- |
| chr1 | 1014 |
| chr2 | 1391 |
| chr3 | 946 |
| chr4 | 815 |
| chr5 | 801 |
| chr6 | 2087 |
| chr7 | 859 |
| chr8 | 771 |
| chr9 | 795 |
| chr10 | 552 |
| chr11 | 694 |
| chr12 | 689 |
| chr13 | 385 |
| chr14 | 257 |
| chr15 | 310 |
| chr16 | 747 |
| chr17 | 1095 |
| chr18 | 240 |
| chr19 | 751 |
| chr20 | 519 |
| chr21 | 153 |
| chr22 | 300 |
| chrX | 67 |

